# “Incidence and Outcomes of Pulmonary embolism among hospitalized COVID-19 patients”

**DOI:** 10.1101/2021.02.16.21251676

**Authors:** Omaima Ibrahim Badr, Hassan Alwafi, Wael Aly Elrefaey, Abdallah Y Naser, Mohammed Shabrawishi

## Abstract

**Background:** Patients with COVID-19 may be at high risk for thrombotic complications due to excess inflammatory response and stasis of blood flow. This study aims to assess the incidence of pulmonary embolism among hospitalized patients with COVID-19, risk factors and the impact on survival.

**Method:** A retrospective case-control study was conducted at Al-Noor Specialist Hospital in, Saudi Arabia between March 15, 2020, and June 15, 2020. Patients with confirmed COVID-19 diagnosis by a real-time polymerase chain reaction (PCR) and confirmed diagnosis of pulmonary embolism by Computed Tomography pulmonary angiogram (CTPA) formed the case group. Patients with confirmed COVID-19 diagnosis by a real-time polymerase chain reaction (PCR) and without confirmed diagnose of pulmonary embolism formed the control group. Logistic regression analysis was used to identify predictors of pulmonary embolism and its survival.

**Results:** A total of 159 patients participated were included in the study, of which 51 were the cases (patients with pulmonary embolism) and 108 patients formed the control group (patients without pulmonary embolism). The incidence of PE among hospitalized was around 32%. Smoking history, low level of oxygen saturation and higher D-dimer values were important risk factors that were associated with a higher risk of developing PE (p< 0.05). Higher respiratory rate was associated with higher odds of death, and decrease the possibility of survival among hospitalised patients with PE.

**Conclusion:** Pulmonary embolism is common among hospitalized patients with COVID-19. Preventive measures should be considered for hospitalized patients with smoking history, low level of oxygen saturation, high D-dimer values, and high respiratory rate.

## Introduction

The novel coronavirus, severe acute respiratory syndrome coronavirus 2 (SARS-CoV-2), emerged in Wuhan city, China at the end of 2019 and since then it has spread in more than 190 countries across the world. The total number of confirmed cases as of August 2020 was □20 million, with □ 700,000 confirmed deaths (1). The severity of the disease can range from being asymptomatic, mild or moderate to severe, with multi-organ failure and death (2, 3). Acute respiratory distress syndrome, which is one of the major complications of coronavirus disease of 2019 (COVID-19), is associated with a high mortality rate and is considered the main cause of death (4, 5). Venous and arterial thrombosis is considered one of the most severe consequences of the disease and has poor prognostic outcomes (6, 7). Patients with COVID-19 may be at a high risk of thrombotic complications due to excess inflammatory response, platelet activation, endothelial dysfunction, and stasis of blood flow (8, 9). At present, it has been suggested that patients with COVID-19 infection have an increased risk of thrombosis (10). In addition, recent data has suggested that increased levels of the D-dimer test can be a predictor of adverse outcomes such as underlying coagulopathy and thrombotic risk in patients with COVID-19 (11, 12). However, the published data are very limited, with no previous studies on the incidence and risk factors of PE in patients with COVID-19 in the Middle East. Therefore, this study aims to assess the incidence of pulmonary embolism among hospitalized patients with COVID-19, risk factors, and the impact on survival.

## METHODS

### Study Design and participants

A retrospective case-control study was conducted at Al-Noor Specialist Hospital in Mecca, Saudi Arabia. The description of the study settings and the hospital has been described previously (1). All patients had a confirmed COVID-19 diagnosis by a real-time polymerase chain reaction (PCR). The PCR samples were obtained through a nasopharyngeal swap. The patients underwent a Computed Tomography pulmonary angiogram (CTPA) for the diagnosis of pulmonary embolism (PE). All patients were admitted between March 15, 2020, and June 15, 2020, and they were followed up for a time to assess the clinical outcome, and the final date of follow-up was August 15, 2020. Patients younger than 15 years old, and patients who were not eligible for CTPA pulmonary angiography, were excluded from the study.

### Data Collection and study variables

Data were collected from medical files and electronic records using a unique medical record number (MRN) for each patient. Data included the patient’s demographics, clinical symptoms (fever, cough, SOB, nausea, vomiting, diarrhoea, headache, body aches, loss of smell and loss of taste), comorbidities and chest radiograph. These data were collected on admission to the hospital. Clinical signs, including heart rate, respiratory rate and SO2% on room air, were collected at the time of CTPA. Data regarding risk assessments for thromboembolism including age, smoking, obesity (defined as BMI □ 30), D-dimer level, the severity of the disease (critical (in ICU) and non-critical (outside ICU)), the time between the admission and the CTPA, the presence of comorbidities, the risk of disseminated intravascular coagulation (DIC), including CBC (WBC, platelets and haemoglobin level), and the coagulation parameters (PT, PTT and INR) were collected in the same day or within 24 hours of the Computed Tomography pulmonary angiogram (CTPA). The radiological examinations including the CTPA were reviewed by a senior certified chest radiologist that classified the pulmonary embolism to the main trunk, lobar, segmental and subsegmental; RV strain by CT was defined as a right ventricle to left ventricle size ratio of ≥ 0.9. In addition, pulmonary systolic pressure, right ventricular (RV) dysfunction (diagnosed by qualitative and quantitative RV dilation or RV systolic dysfunction), left ventricular (LV) dysfunction (defined as decreased left ventricular systolic function with decreased LV ejection fraction, regional wall motion abnormalities or both), and visualization of thrombus in the right heart or the pulmonary artery were diagnosed through trans-thoracic echocardiography, which was reviewed by an experienced, board-certified echocardiography attending physician.

### Case definition

Patients with a confirmed COVID-19 diagnosis by a real-time polymerase chain reaction (PCR) and without a confirmed diagnosis of pulmonary embolism formed the control group.

### Control definition

Patients with a confirmed COVID-19 diagnosis by a real-time polymerase chain reaction (PCR) and without a confirmed diagnosis of pulmonary embolism formed the control group.

### Outcomes

The outcome predictors of those patients were admission to an intensive care unit, intubation and connection to mechanical ventilation, thrombolytic therapy or catheter-directed thrombolysis, bleeding (minor or major bleeding) as a complication of PE therapy, a prolonged hospital stay (□ 2 weeks), and mortality.

### Statistical Analysis

Descriptive statistics were used to describe patients’ demographic characteristics, radiological findings, clinical signs and symptoms, and comorbidities. Continuous data were reported as mean ± SD for normally distributed data and as median (interquartile range (IQR)) for not normally distributed data, and categorical data were reported as percentages (frequencies). Independent sample t-test/ANOVA was used to compare the mean value for normally distributed continuous variables and The Mann-Whitney U test/Kruskal-Wallis test was used to compare the median value for not normally distributed ones. A Chi-squared test/Fisher test was used as appropriate to compare proportions for categorical variables. Logistic regression analysis was used to identify predictors of pulmonary embolism and its survival. A confidence interval of 95% (p < 0.05) was applied to represent the statistical significance of the results, and the level of significance was assigned as 5%. SPSS (Statistical Package for the Social Sciences) version 25.0 software (SPSS Inc.) was used to perform all statistical analysis.

### Ethical approval

This study was approved by the institutional ethics board at the Ministry of Health in Saudi Arabia (No H-02-K-076-0920-386).

### Patient and public involvement

It was not appropriate or possible to involve patients or the public in the design, or conduct, or reporting, or dissemination plans of our research.

## Results

### Patient’s baseline characteristics

A total of 159 patients participated in the study, of which 51 were the cases (patients with pulmonary embolism) and 108 patients formed the control group (patients without pulmonary embolism). The incidence of PE among hospitalized patients was around 32%. The case and control group showed comparable baseline characteristics; there was no statistically significant difference between the two groups in term of the gender distribution, duration of stay at the hospital, disease history, and most of the reported signs and symptoms (p> 0.05). For further details on the baseline characteristics, refer to **Table 1** below.

**Table 1:** Baseline characteristics for the study sample.

| Demographic variable | Patients with Pulmonary embolism (case) (n= 51) | Patients without Pulmonary embolism (control) (n= 108) | P-Value |
| --- | --- | --- | --- |
| **Demographics** |  |  |  |
| Age (years) | 56.9 (12.3) | 50.9 (15.2) | 0.009** |
| Gender No. (%) |  |  |  |
| Male | 40 (78.4) | 71 (65.7%) | 0.139 |
| BMI (kg/m2) | 27.7 (5.8) | 27.9 (5.8) | 0.609 |
| **Smoking history** |  |  |  |
| Smoker | 32 (62.7) | 29 (26.9) | 0.000*** |
| **Comorbidities No. (%)** |  |  |  |
| Diabetes mellitus | 24 (47.1) | 43 (39.8) | 0.388 |
| Hypertension | 21 (41.2) | 38 (35.2) | 0.465 |
| Ischemic heart disease | 10 (19.6) | 17 (15.7) | 0.544 |
| Heart failure | 3 (5.9) | 6 (5.6) | 0.934 |
| Renal failure | 0 | 7 (6.5) | 0.060 |
| Malignancy | 1 (2.0) | 0 | 0.144 |
| HIV | 0 | 1 (0.9) | 0.491 |
| **Rheumatologically disease No. (%)** |  |  |  |
| Rheumatoid arthritis | 0 | 1 (0.9) | 0.491 |
| Antiphospholipid syndrome | 0 | 1 (0.9) | 0.491 |
| **Haematological disease No. (%)** |  |  |  |
| Sickle cell disease | 0 | 3 (2.8) | 0.230 |
| **Pulmonary disease (other than pulmonary embolism) No. (%)** |  |  |  |
| COPD | 1 (2.0) | 7 (6.5) | 0.854 |
| Asthma | 1 (2.0) | 8 (7.4) | 0.719 |
| Tuberculosis | 0 | 2 (1.9) | 0.544 |
| IPF | 0 | 1 (0.9) | 0.676 |
| Pulmonary hypertension | 1 (2.0) | 0 | 0.012* |
| **Sign and symptoms (at presentation to hospital) No. (%)** |  |  |  |
| Fever | 44 (86.3) | 80 (74.1) | 0.083 |
| Cough | 41 (80.4) | 85 (78.7) | 0.806 |
| Sore throat | 20 (39.2) | 47 (43.5) | 0.608 |
| Dyspnoea | 47 (92.2) | 98 (90.7) | 0.769 |
| Haemoptysis | 7 (13.5) | 11 (10.2) | 0.511 |
| Chest pain | 21 (41.2) | 26 (24.1) | 0.027* |
| Vomiting | 6 (11.8) | 28 (25.9) | 0.042* |
| Diarrhea | 15 (29.4) | 33 (30.6) | 0.883 |
| Nausea | 21 (41.2) | 39 (36.1) | 0.539 |
| Loss of smell | 4 (7.8) | 18 (16.7) | 0.133 |
| Loss of taste | 4 (7.8) | 17 (15.7) | 0.170 |
| Headache | 22 (43.1) | 42 (38.9) | 0.610 |
| Bone ache | 29 (56.9) | 64 (59.3) | 0.775 |
| **Other No. (%)** |  |  |  |
| Duration of stay at hospital (median days (IQR)) | 13.00 (13.00) | 15.00 (11.00) | 0.983 |
| Duration between admission and spiral (median days) | 6.00 (7.00) | 5.00 (6.75) | 0.953 |
| **Outcomes No. (%)** |  |  |  |
| Survived | 38 (74.5) | 74 (68.5) | 0.440 |
| Died | 13 (25.5) | 34 (31.5) | 0.440 |
| ICU admission (yes) | 39 (76.5) | 80 (74.1) | 0.745 |
| Intubation (yes) | 60 (55.6) | 32 (62.7) | 0.391 |
| Duration of stay at the ICU (median days (IQR)) | 6.00 (10.00) | 7.00 (13.00) | 0.524 |
| **Medications and management No. (%)** |  |  |  |
| Mechanical thrombectomy | 3 (5.9) | 0 | 0.803 |
| Alteplase | 1 (2.0) | 0 | 0.888 |
| **Complications of anticoagulant No. (%)** |  |  |  |
| Minor bleeding | 3 (5.9) | 0 | 0.011* |

### Vital signs and blood test findings

The case and control groups were comparable in terms of their vital sign measurements (respiratory rate, heart rate, oxygen saturation) and various blood test measurements (haematocrit, HGB, WBC, INR, and PTT). However, PT and D-dimer measures were significantly different between the two groups, where the mean PT value was higher in the control group compared to the case group, and the median D-dimer value was higher among the case group compared to the control group. For further details on vital signs and blood test findings, refer to **Table 2**.

**Table 2:**
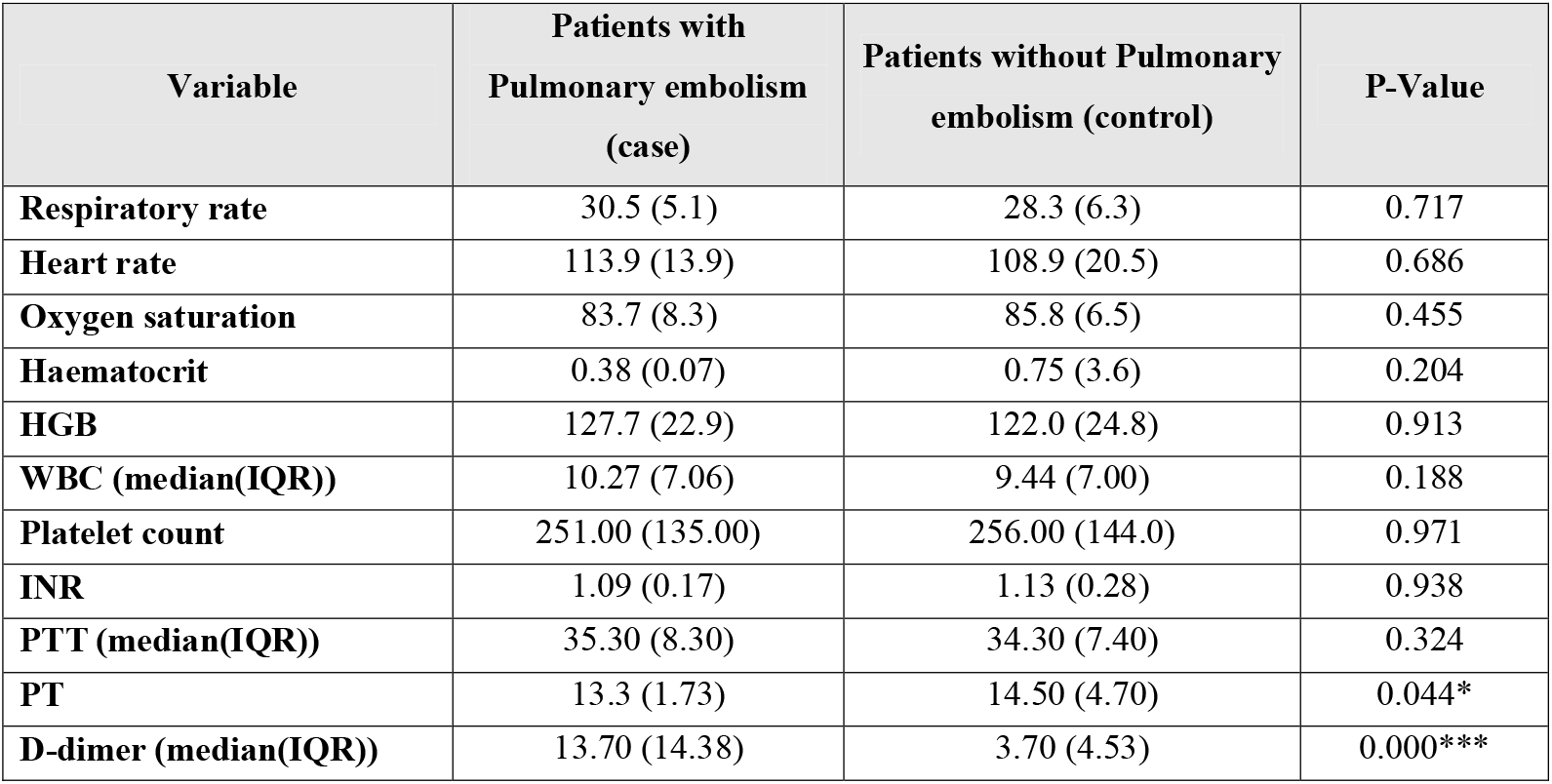
Vital signs and blood tests upon admission.

| Variable | Patients with<br>Pulmonary embolism<br>(case) | Patients without Pulmonary<br>embolism (control) | P-Value |
| --- | --- | --- | --- |
| **Respiratory rate** | 30.5 (5.1) | 28.3 (6.3) | 0.717 |
| **Heart rate** | 113.9 (13.9) | 108.9 (20.5) | 0.686 |
| **Oxygen saturation** | 83.7 (8.3) | 85.8 (6.5) | 0.455 |
| **Haematocrit** | 0.38 (0.07) | 0.75 (3.6) | 0.204 |
| **HGB** | 127.7 (22.9) | 122.0 (24.8) | 0.913 |
| **WBC (median(IQR))** | 10.27 (7.06) | 9.44 (7.00) | 0.188 |
| **Platelet count** | 251.00 (135.00) | 256.00 (144.0) | 0.971 |
| **INR** | 1.09 (0.17) | 1.13 (0.28) | 0.938 |
| **PTT (median(IQR))** | 35.30 (8.30) | 34.30 (7.40) | 0.324 |
| **PT** | 13.3 (1.73) | 14.50 (4.70) | 0.044* |
| **D-dimer (median(IQR))** | 13.70 (14.38) | 3.70 (4.53) | 0.000*** |

### Patient’s radiological findings

The majority of pulmonary emboli were located in the segmental branch. There was no statistically significant difference between the case and the control groups in terms of the CT parenchymal findings (p> 0.05). RT ventricular dysfunction was more common among the case group compared to the control group, whereas the distribution of LT ventricular dysfunction and systolic pulmonary embolism between the two groups was the same **(Table 3)**.

**Table 3:** Radiological and ECHO findings.

| Variable | Patients with Pulmonary embolism (case) | Patients without Pulmonary embolism (control) | P-value |
| --- | --- | --- | --- |
| **Location of pulmonary embolism** |  |  |  |
| Segmental branch | 23 (45.1) | - | - |
| Main pulmonary trunk | 12 (23.5) | - | - |
| Lobar branch | 10 (19.6) | - | - |
| Sub segmental | 6 (11.8) | - | - |
| **CT parenchymal findings** |  |  |  |
| Bilateral peripheral ground glass | 26 (51.0) | 50 (46.3) | 0.581 |
| Bilateral peripheral ground glass with | 23 (45.1) | 50 (46.3) | 0.887 |
| Unilateral peripheral ground glass | 2 (3.9) | 8 (7.4) | 0.398 |
| **ECHO** |  |  |  |
| *RT ventricular dysfunction* | 25 (49.0) | 22 (20.4) | 0.000*** |
| *LT ventricular dysfunction* | 4 (7.8) | 8 (7.4) | 0.575 |
| *Systolic pulmonary hypertension* | 14 (27.5) | 24 (22.2) | 0.298 |

### Risk factors of pulmonary embolism and its survival

Using unadjusted logistic regression, we identified that age, smoking history, a higher respiratory rate, and a higher D-dimer value were associated with a higher risk of developing PE (p< 0.05). A higher respiratory rate, higher WBC, higher INR value, longer duration between admission and spiral, longer duration of stay at the hospital, requiring ICU admission, duration of stay at the ICU, and requiring intubation were associated with higher odds of death, and a decrease in the possibility of survival among hospitalized patients with PE (p< 0.05). On the other hand, a higher HGB value was associated with higher odds of survival (p< 0.05).

Adjusting for relevant confounding variables such as age, gender, BMI, renal failure, malignancy, haematological diseases, duration between admission and spiral, duration in the ICU, and intubation, we identified that smoking history, a low level of oxygen saturation, and higher D-dimer values were important risk factors that were associated with a higher risk of developing PE (p< 0.05). A higher respiratory rate was associated with higher odds of death, and a decrease in the possibility of survival among hospitalized patients with PE. For further details on the findings of the logistic regression analysis, refer to **Table 4**.

**Table 4:** Risk factors of pulmonary embolism and its survival.

| Variable | Risk factors of pulmonary embolism |  |  |  | Factors affecting survival among patients with pulmonary embolism |  |  |  |
| --- | --- | --- | --- | --- | --- | --- | --- | --- |
|  | OR (95%CI) | P-value | AOR§ (95%CI) | P-value | OR (95%CI) | P-value | AOR§ (95%CI) | P-value |
| **Demographics and social** |  |  |  |  |  |  |  |  |
| Age (years) | 1.03 (1.01 – 1.06) | **0.018\*** | - |  | 0.99 (0.96 – 1.01) | 0.230 | - |  |
| Gender (female) | 0.53 (0.24 – 1.15) | 0.107 | - |  | 1.19 (0.56 – 2.53) | 0.653 | - |  |
| Smoking (yes) | 4.59 (2.26 – 9.33) | **0.000\*\*\*** | 3.85 (1.73 – 8.58) | **0.001\*\*** | 1.00 (0.50 – 2.02) | 0.991 | 1.70 (0.46 – 6.27) | 0.422 |
| Obesity (BMI> 30 kg/m<sup>2</sup>) | 1.40 (0.70 – 2.81) | 0.337 | - |  | 0.87 (0.43 – 1.78) | 0.703 | - |  |
| **Comorbidities** |  |  |  |  |  |  |  |  |
| Diabetes mellitus (yes) | 1.34 (0.69 – 2.63) | 0.388 | 1.02 (0.47 – 2.19) | 0.969 | 0.76 (0.38 – 1.52) | 0.440 | 1.14 (0.35 – 3.71) | 0.833 |
| Hypertension (yes) | 1.29 (0.65 – 2.55) | 0.466 | 1.21 (0.56 – 2.61) | 0.623 | 0.64 (0.32 – 1.28) | 0.202 | 0.65 (0.20 – 2.09) | 0.473 |
| Ischemic heart disease (yes) | 1.31 (0.55 – 3.10) | 0.545 | 1.05 (0.40 – 2.77) | 0.915 | 0.81 (0.33 – 1.96) | 0.638 | 0.88 (0.20 – 3.87) | 0.870 |
| Heart failure (yes) | 1.06 (0.26 – 4.43) | 0.934 | 1.22 (0.25 – 6.09) | 0.806 | 0.50 (0.13 – 1.96) | 0.322 | 0.05 (0.00 – 1.24) | 0.068 |
| Renal failure (yes) | - |  | - |  | - |  | - |  |
| Malignancy (yes) | - |  | - |  | - |  | - |  |
| Rheumatologically disease (yes) | - |  | - |  | - |  | - |  |
| Haematological disease (yes) | - |  | - |  | - |  | - |  |
| COPD (yes) | 0.79 (0.06 – 10.38) | 0.855 | - |  | 0.90 (0.12 – 7.03) | 0.920 | - |  |
| Asthma (yes) | 0.63 (0.05 – 8.20) | 0.720 | - |  | 1.17 (0.15 – 9.01) | 0.882 | - |  |
| Tuberculosis (yes) | - |  | - |  | - |  | - |  |
| IPF (yes) | - |  | - |  | - |  | - |  |
| Pulmonary hypertension (yes) | - |  | - |  | - |  | - |  |
| **Vital signs** |  |  |  |  |  |  |  |  |
| **Respiratory rate** | 1.07 (1.00 – 1.15) | **0.043\*** | 1.07 (0.99 – 1.15) | 0.076 | 0.91 (0.84 – 0.98) | **0.011\*** | 0.82 (0.71 – 0.94) | **0.004\*\*** |
| **Heart rate** | 1.02 (1.00 – 1.04) | 0.067 | 1.02 (1.00 – 1.05) | 0.061 | 0.98 (0.96 – 1.00) | 0.113 | 0.97 (0.94 – 1.01) | 0.165 |
| **Oxygen saturation** | 0.98 (0.95 – 1.02) | 0.279 | 0.95 (0.90 – 1.00) | **0.039\*** | 1.02 (0.99 – 1.06) | 0.272 | 1.05 (0.97 – 1.15) | 0.230 |
| **Laboratory** |  |  |  |  |  |  |  |  |
| **Haematocrit** | 0.89 (0.48 – 1.64) | 0.705 | 0.74 (0.01 – 65.73) | 0.895 | - |  | - |  |
| **HGB** | 1.01 (0.99 – 1.02) | 0.438 | 1.00 (0.98 – 1.02) | 0.790 | 1.03 (1.01 – 1.04) | **0.001\*\*** | 1.01 (0.98 – 1.03) | 0.528 |
| **WBC** | 1.02 (0.97 – 1.07) | 0.558 | 1.02 (0.97 – 1.08) | 0.407 | 0.94 (0.89 – 1.00) | **0.035\*** | 0.97 (0.88 – 1.06) | 0.468 |
| **Platelet count** | 1.00 (1.00 – 1.00) | 0.615 | 1.00 (1.00 – 1.00) | 0.972 | 1.00 (1.00 – 1.00) | 0.580 | 1.00 (1.00 – 1.00) | 0.819 |
| **PT** | 0.91 (0.80 – 1.04) | 0.174 | 0.91 (0.80 – 1.04) | 0.180 | 1.04 (0.94 – 1.16) | 0.434 | 1.00 (0.813 – 1.23) | 0.985 |
| **PTT** | 1.00 (0.99 – 1.01) | 0.642 | 1.00 (0.99 – 1.01) | 0.491 | 1.00 (1.00 – 1.01) | 0.645 | 1.03 (0.97 – 1.09) | 0.310 |
| **INR** | 1.07 (0.26 – 4.34) | 0.925 | 2.31 (0.30 – 17.65) | 0.419 | 0.12 (0.02 – 0.69) | **0.018\*** | 0.05 (0.00 – 1.20) | 0.065 |
| **D dimer** | 1.17 (1.10 – 1.24) | **0.000\*\*\*** | 1.21 (1.13 – 1.30) | **0.000\*\*\*** | 0.99 (0.96 – 1.01) | 0.356 | 0.99 (0.95 – 1.03) | 0.685 |
| **Other** |  |  |  |  |  |  |  |  |
| **Duration between admission and spiral (days)** | 0.99 (0.92 – 1.07) | 0.862 | - |  | 0.89 (0.83 – 0.96) | **0.003\*\*** | - |  |
| **Severity of disease (required ICU admission)** | 1.14 (0.52 – 2.47) | 0.745 | 1.27 (0.46 – 3.48) | 0.646 | 0.04 (0.01 – 0.31) | **0.002\*\*** | 1.79 (0.09 – 36.12) | 0.705 |
| **Duration of hospital stay (days)** | 1.00 (0.96 – 1.04) | 0.907 | 1.01 (0.96 – 1.07) | 0.695 | 0.96 (0.93 – 1.00) | **0.042\*** | 1.10 (1.00 – 1.21) | 0.053 |
| **Duration of stay in the ICU (days)** | 0.98 (0.94 – 1.02) | 0.395 | - |  | 0.86 (0.82 – 0.91) | **0.000\*\*\*** | - |  |
| **Intubation (yes)** | 0.74 (0.38 – 1.47) | 0.392 | - |  | 0.01 (0.00 – 0.05) | **0.000\*\*\*** | - |  |

§ Adjusted for the following variables: age, gender, BMI, renal failure, malignancy, haematological diseases, duration between admission and spiral, duration in the ICU, and intubation

## Discussion

To the best of our knowledge, this is the first study that investigated the incidence and risk factors of pulmonary embolism among hospitalized patients with COVID-19 in the Middle East. In this case-control study, we investigated the incidence, risk factors and clinical outcomes among hospitalized patients with pulmonary embolism and COVID-19. The key findings of this study are that smoking history, a low level of oxygen saturation, and higher D-dimer values were important risk factors that were associated with a higher risk of developing PE (p< 0.05). A higher respiratory rate was associated with higher odds of death and a decrease in the possibility of survival among hospitalized patients with PE. We found that around 32% of hospitalized patients with COVID-19 who underwent CTPA were diagnosed with PE. This finding coincides with previous studies, which reported that about 37.1% of COVID-19 patients were diagnosed with PE (2). Patients with smoking history values were associated with a higher risk of developing PE. Previous reviews reported a higher risk in current smokers than in individuals who have never smoked (3). Smoking may promote VTE via different mechanisms such as a procoagulant state, reduced fibrinolysis, inflammation, and increased blood viscosity (4, 5).

In our study, elevated D-dimer was found in all patients with PE and non-PE; an elevated D-dimer was found in other reports on COVID −19 associated with pneumonia. Researchers attributed this elevation to excessive consumption of coagulation factors. Some other studies showed that up to 90% of patients admitted to the hospital for pneumonia had high procoagulant markers, with D-dimer being one of the most common (6). In our study, there is a significant elevation of D-dimer in the PE group compared to the control group, indicating that CTPA is necessary for the exclusion of PE if there is clinical deterioration. This finding coincides with previous studies, which reported thatelevated D-dimer levels correlate with the presence of VTE in COVID-19 patients (7, 8). In our study, patients without PE had a median D-dimer level of 3.7, similar to 3.3 observed in patients with Covid-19 in the study by Reich et al. (9).

Pharmacological thromboprophylaxis has shown promise in preventing venous thromboembolism (VTE) for high-risk individuals. The incidence of VTE ranges from 5% to 15% and can be effectively reduced by one-half to two-thirds with appropriate thromboprophylaxis (10). In critically ill patients, the incidence of deep vein thrombosis (DVT) ranges from 13% to 31% without thromboprophylaxis (11). However, this risk can be reduced with pharmacological thromboprophylaxis (12). In our study, all patients were on thromboprophylaxis per guidelines during hospitalization. This finding heightened the risk of venous thromboembolism in COVID-19, despite prophylactic anticoagulation.

The association between COVID-19 and hypercoagulopathy had been established in the literature. This hypercoagulable state is found to be correlated with the severity of the illness since both the incidence and the coagulation abnormalities are more apparent in severe cases. Furthermore, the presence of coagulation abnormalities such as elevated D-dimer has been linked with both mortality and the need for mechanical ventilation (13). Despite receiving anticoagulation with at least one prophylactic dose, the incidence of VTE was approximately 24% (8). Several mechanisms contributed to the pathogenesis of micro and macro thrombotic complications in COVID-19. These include the systemic activation of coagulation, and, like other viruses (SARS-COV1 and MERS-COV), SARS-COV2 promotes endothelial dysfunction, vascular leak, and pulmonary microthrombi (14-16).

The major entry receptors of SARS-CoV-2, the angiotensin converting enzyme 2 (ACE2), are located at the cell surfaces of many organs and regulate renin angiotensin system (RAS). This binding results in dysregulation of RAS and an increase in the pro-inflammatory cytokines ensuing endothelial dysfunction and micro thrombosis (17-19). Autopsy studies of COVID-19 cases have shown that venous thromboembolic events (PE or DVT) are common. Additionally, features of micro thrombosis and endotheliitis were reported not only in the lungs (20, 21) but also in the heart, intestine and kidneys (22). Additional mechanisms were also described as antiphospholipid antibodies (APSA), where recent publications have reported the association of APSA with thrombotic complications observed in COVID-19 patients (23).

We did not find significant differences between the PE and non-PE group regarding ICU admission, connection to mechanical ventilation, survival, or duration of hospital stay. However, the mortality in patients with PE was around 25.5%; this finding coincides with previous studies, which reported a mortality rate of about 20% (24). Also, we found no differences between both groups in the other risk factors for PE, in the duration of hospital admission, and in the duration between admission and the time of CTPA between both groups; however, this may be due to up-regulation of procoagulant activity in COVID-19 infection increasing the risk of PE. The coexistence of pneumonia and PE has been known for years, and data from the international cohort RIETE showed that patients with respiratory infections had a higher risk of PE procoagulant activity than patients with other types of infections (25). Patients with coronavirus disease 2019 (COVID-19) pneumonia may be predisposed to thrombotic complications due to excess inflammatory response, endothelial dysfunction, platelet activation, and stasis of blood flow (26). In our study, there was no significant difference between both groups in the duration of hospital admission or the duration between admission and the time of CTPA, suggesting that there are other mechanisms related to COVID-19 that promote the coagulopathy, which should be studied in the future.

The results of this study demonstrated a high incidence of pulmonary embolism among hospitalized patients with COVID-19, and this finding should alert health care providers to suspect a diagnosis of PE in COVID-19 patients, especially when clinical signs are associated with significant elevation of D-dimer. Further studies are required to determine the duration of thromboprophylaxis in COVID-19 patients.

This study has some limitations. First, the number of patients included in the study was small, and the study population only included patients from a single-centre hospital in Saudi Arabia. Second, due to the retrospective nature of the study, we did not have data on the CTPA for all suspected PE patients who were limited by clinical instability or a refusal to do CT with IV contrast. In addition, we were not able to determine if the PE occurred in hospital or before hospital admission.

## Conclusions

Our results revealed a high incidence of pulmonary embolism among hospitalized patients with COVID-19. Preventive measures should be considered for hospitalized patients with smoking history, low level of oxygen saturation, high D-dimer values, and a high respiratory rate. More studies are required to evaluate the risk factors and the mechanisms of thrombosis related to COVID-19.

## AUTHORS CONTRIBUTIONS

Conceptualization, Hassan Alwafi, Omaima Badr and Wael Elrefaey; Data curation, Omaima Badr and Wael Elrefaey; Formal analysis, Abdallah Y Naser and Hassan Alwafi; Investigation, Omaima Badr and Hassan Alwafi; Methodology, Abdallah Y Naser and Hassan Alwafi; Project administration, Omaima Badr and Hassan Alwafi; Resources, Mohammed Shabrawishi, Omaima Badr, Hassan Alwafi and Wael Elrefaey; Supervision, Omaima Badr and Hassan Alwafi; Validation, Omaima Badr and Hassan Alwafi; Writing – original draft, Mohammed Shabrawishi, Abdallah Y Naser, Omaima Badr and Hassan Alwafi; Writing – review & editing, Mohammed Shabrawishi, Abdallah Y Naser, Omaima Badr, Wael Elrefaey and Hassan Alwafi

## Data Availability

No further data is available

## CONFLICT OF INTEREST

The authors have stated explicitly that there are no conflicts of interest in connection with this article.

## Notes

### Competing Interest Statement

The authors have declared no competing interest.

### Funding Statement

No fund

### Author Declarations

This study was approved by the institutional ethics board at the Ministry of Health 137 in Saudi Arabia (No H-02-K-076-0920-386

